# An improved mathematical prediction of the time evolution of the Covid-19 Pandemic in Italy, with Monte Carlo simulations and error analyses

**DOI:** 10.1101/2020.04.20.20073155

**Authors:** Ignazio Ciufolini, Antonio Paolozzi

**Affiliations:** Dipartimento di Ingegneria dell’Innovazione, University of Salento, Lecce and Centro Fermi, Rome, Italy; Scuola di Ingegneria Aerospaziale, Sapienza, University of Rome

## Abstract

We present an improved mathematical analysis of the time evolution of the Covid-19 pandemic in Italy and a statistical error analyses of its evolution, including Monte Carlo simulations with a very large number of runs to evaluate the uncertainties in its evolution. A previous analysis was based on the assumption that the number of nasopharyngeal swabs would be constant. However the number of daily swabs is now more than five times what it was when we did our previous analysis. Therefore, here we consider the time evolution of the ratio of the new daily cases to number of swabs, which is more representative of the evolution of the pandemic when the number of swabs is increasing or changing in time. We consider a number of possible distributions representing the evolution of the pandemic in Italy and we test their prediction capability over a period of up to four weeks. The results show that a distribution of the type of Planck black body radiation law provides very good forecasting. The use of different distributions provides an independent possible estimate of the uncertainty. We then consider five possible trajectories for the number of daily swabs and we estimate the potential dates of a substantial reduction in the number of new daily cases. We then estimate the spread in a substantial reduction, below a certain threshold, of the daily cases per swab among the Italian regions. We finally perform Monte Carlo simulations with 25000 runs to evaluate a random uncertainty in the prediction of the date of a substantial reduction in the number of diagnosed daily cases per swab.

## 1. Introduction

In a previous paper, we estimated the possible dates of a substantial reduction in the daily number of new cases of the Covid-19 pandemic based on the assumption that the number of nasopharyngeal swabs would remain roughly constant^1,2^. At the time of our previous analysis (March 26), the average daily number of swabs from February 25 was about 9000 per day. However, from March 27 up to May 16, the average number of daily swabs was about 50600 (recently on May 1, the number of daily swabs reached the maximum of 74208). Therefore, to study the evolution of the Covid-19 pandemic, we have to consider the analysis of the ratio of daily cases per swab. To possibly mathematically predict the evolution of the pandemic in Italy we can fit the ratio of cases per swab using several different distributions. Some distributions are more suitable than others for forecasting the future behavior of the pandemic. We consider the following distributions: Gauss^3^, Beta^3^, Gamma^3^, Weibull^4^, Lognormal^5^ and two versions of the Planck black body radiation law^6^. The number of parameters chosen in these distributions is either two or three, depending on the distribution. It turns out that a Planck law distribution with three independent parameters, along with the Gamma distribution with three parameters, provide the best fits and best predictions.

Furthermore, since the number of daily swabs depends on factors that are unknown to us, such as the daily availability of reagents and specialized personnel, we consider five possible trajectories for the daily number of swabs. (We have also considered some other time evolutions in the number of daily swabs, which for brevity we do not report here.) We fit the time evolution of the cases per unit swab up to April 25, using the two best-fit-prediction distributions, i.e., Planck with three parameters and Gamma, along with a Gauss distribution. After analyzing the time evolution of the cases per unit swab using these three distributions and five conceivable trajectories of daily swabs, we estimate the evolution in the number of new daily cases and the dates of a substantial reduction in such a daily number. Furthermore, in section 3.1 we estimate the spread in the dates of a substantial reduction in the number of daily cases per swab among the regions of Italy, where the conditions are quite different from each other, including the number of swabs per person.

The different distributions that we use provide a possible independent way to estimate the uncertainty. Indeed, a basic problem is to mathematically estimate the uncertainty in the date of a substantial reduction of daily cases. For the purpose of estimating the random uncertainties, in section 3 we report the results of a Monte Carlo simulation with 25000 runs.

## 2 A mathematical analysis of the ratio of new daily cases per swab

After analyzing the time trend of the ratio of new daily cases to the number of daily swabs^7-9^, we found^1,2^ that this trend can be modeled by a Gauss distribution, however the time trend has also a certain amount of skewness that can better be fitted by choosing a skewed distribution such as the Weibull, Log-normal, Beta and Gamma distributions, and also other distributions of the type of the Planck black body law. This last one with three parameters, reported in Fig. 1, shows the best fit and the best prediction capabilities. In Figs. 2a to 2e are reported the fits of the data with a distributions of the type of the Planck black body law with two parameters (i.e., with the exponent of the time variable *t* equal to 3 as in the Planck black body law), and with distributions of the type of the Gamma, Beta, Weibull, Lognormal respectively. The data can also be approximated by a function of the type of a Gauss function with three parameters, as shown in Fig. 2f.

**Fig. 1.**
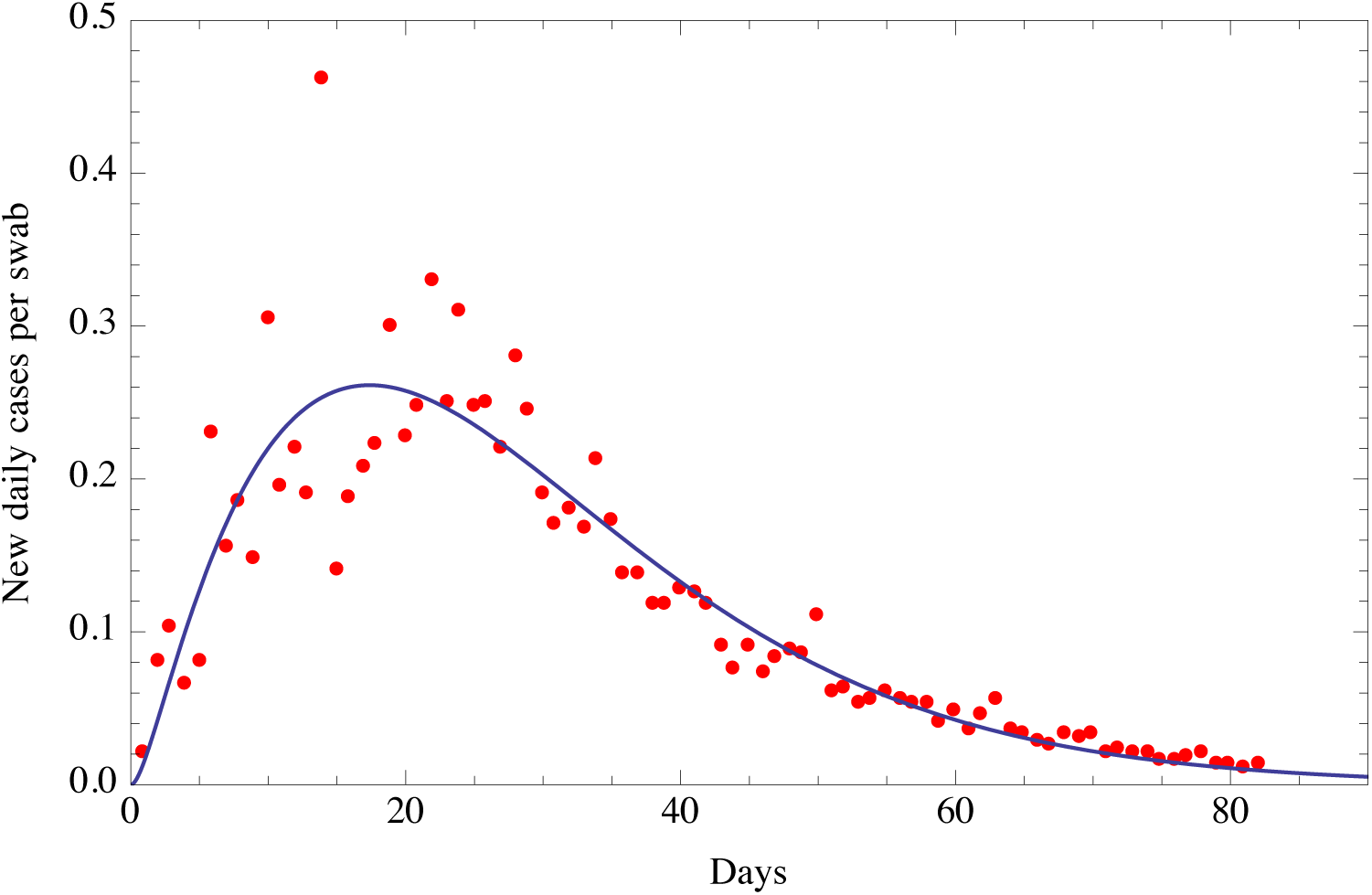
Fit of the ratio of the number of new daily Covid-19 detections in Italy via nasopharyngeal swab tests on a given day divided by the number of nasopharyngeal swab tests given in Italy that day. This fit is obtained with a function of the type of the Planck black body law^6^, 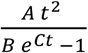, with three parameters *A, B* and *C*. The beginning date is February 25. Root Mean Square of the residuals is 0.0389

**Fig. 2a.**
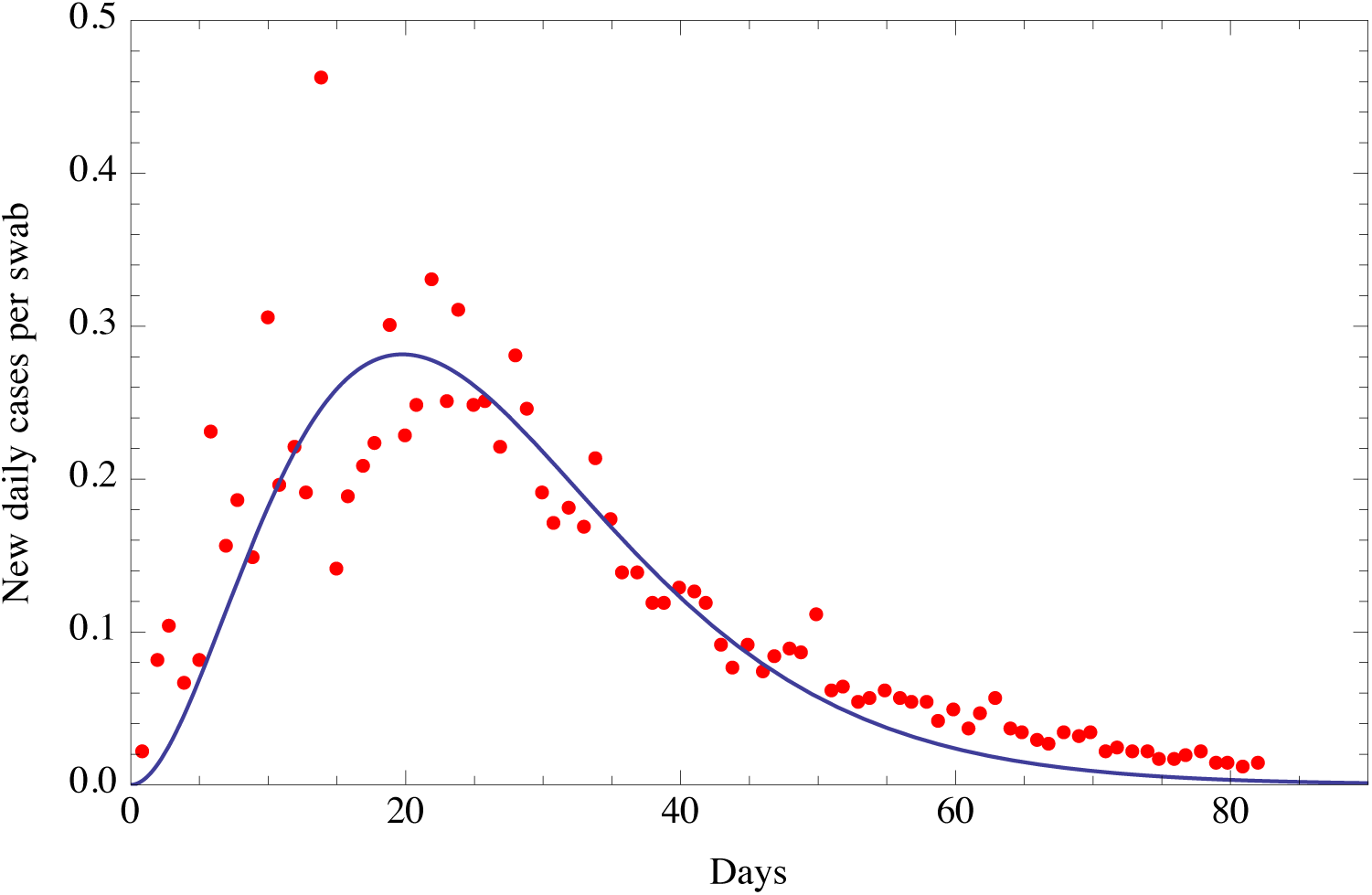
Fit of the ratio of the number of new daily Covid-19 detections in Italy via nasopharyngeal swab tests on a given day divided by the number of nasopharyngeal swab tests given in Italy that day. This fit is obtained with a function of the type of the Planck black body law ^6^, 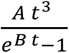, with two parameters *A* and *B*, with the exponent of the time variable *t* equal to 3 as in the Planck law. The beginning date is February 25. Root Mean Square of the residuals is 0.0438

**Fig. 2b.**
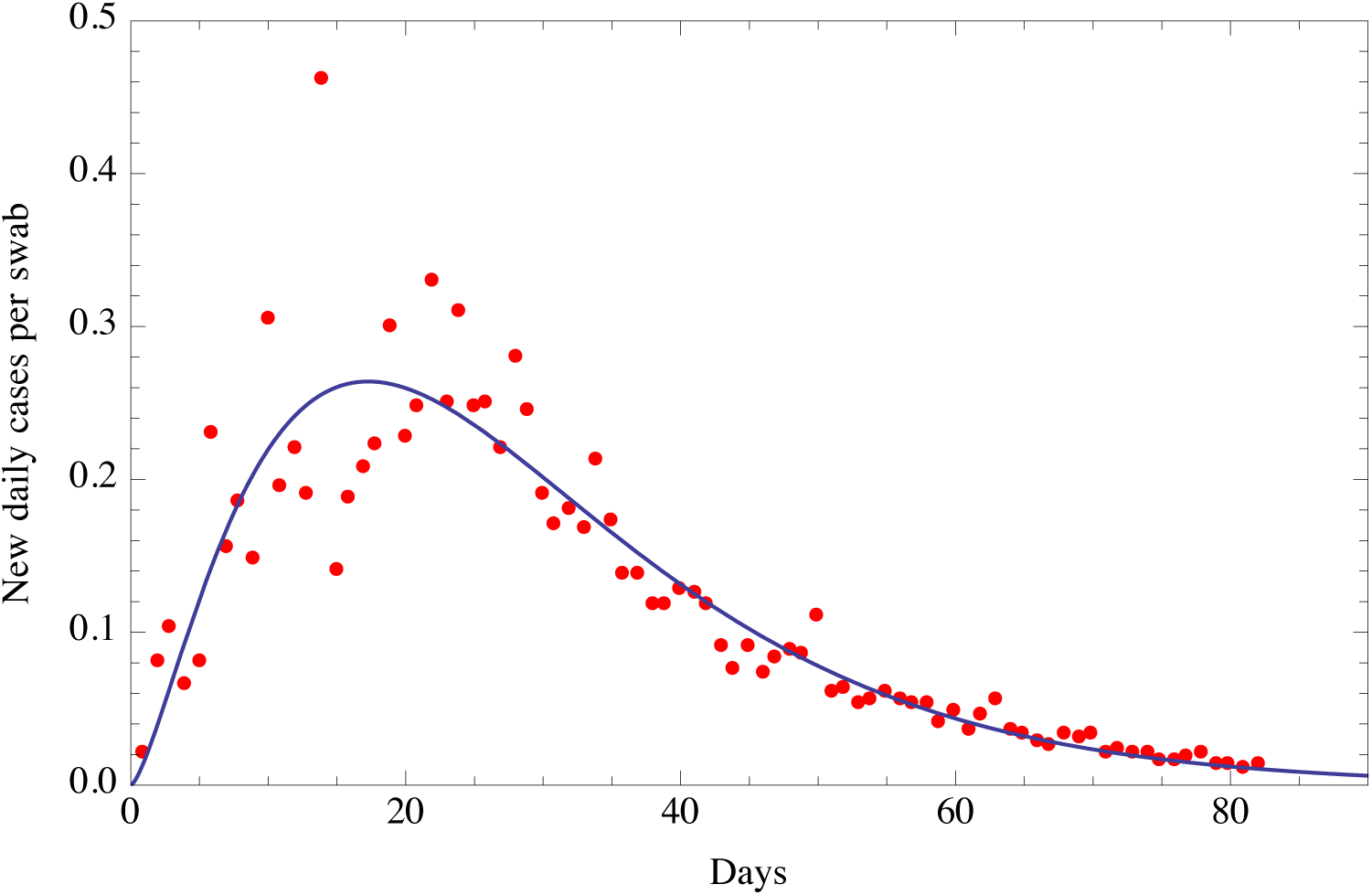
Fit of the ratio of the number of new daily Covid-19 detections in Italy via nasopharyngeal swab tests on a given day divided by the number of nasopharyngeal swab tests given in Italy that day. This fit is obtained with a function of the type of a Gamma distribution^3^, 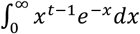, with three parameters *A, B* and *C*, where: 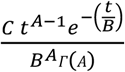. The beginning date is February 25. Root Mean Square of the residuals is 0.0390

**Fig. 2c.**
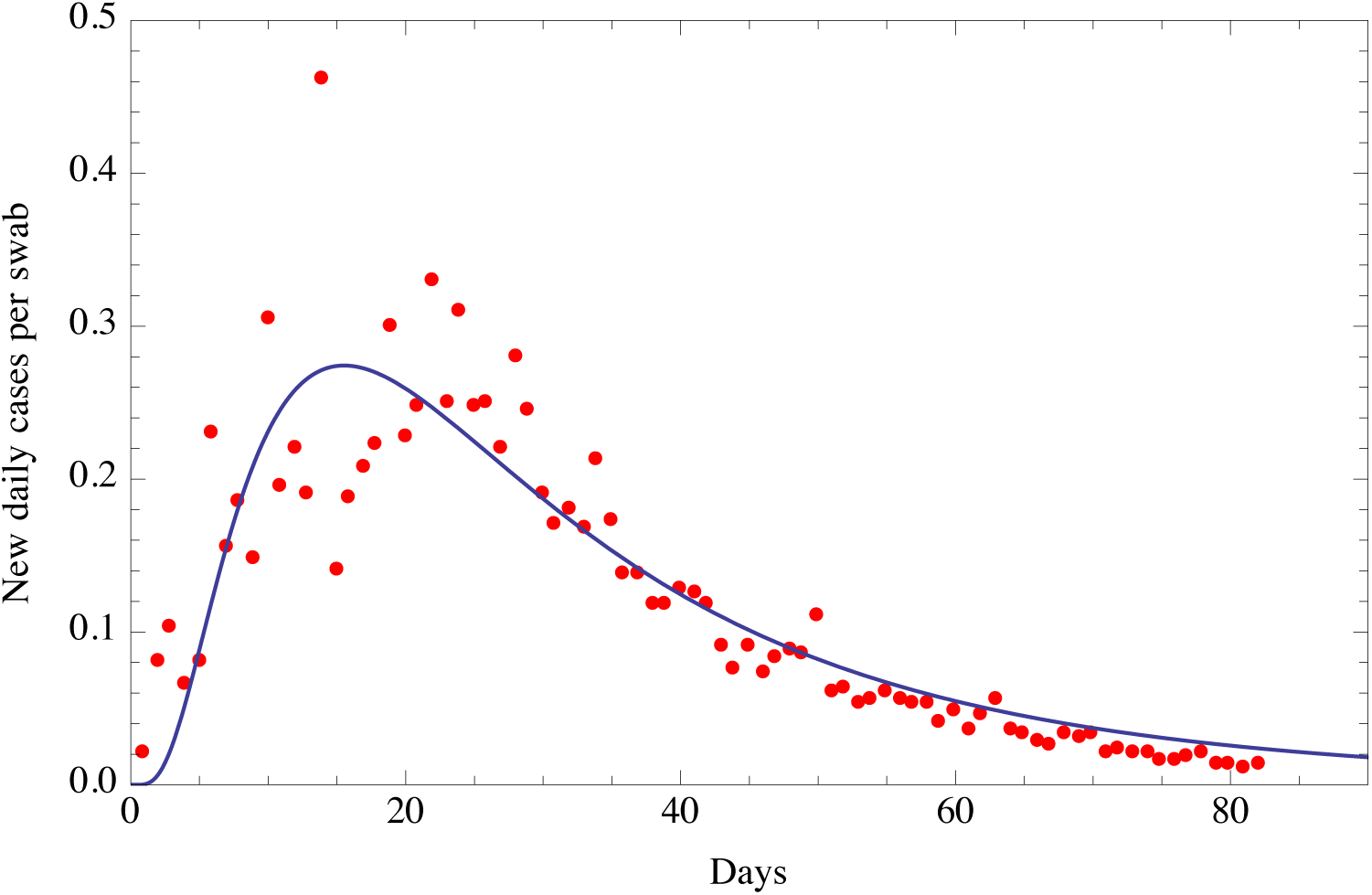
Fit of the ratio of the number of new daily Covid-19 detections in Italy via nasopharyngeal swab tests on a given day divided by the number of nasopharyngeal swab tests given in Italy that day. This fit is obtained with a function of the type of a Beta distribution^3^,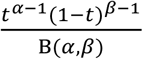, with two parameters *α* and *β*, where: 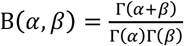. The beginning date is February 25. Root Mean Square of the residuals is 0.0401

**Fig. 2d.**
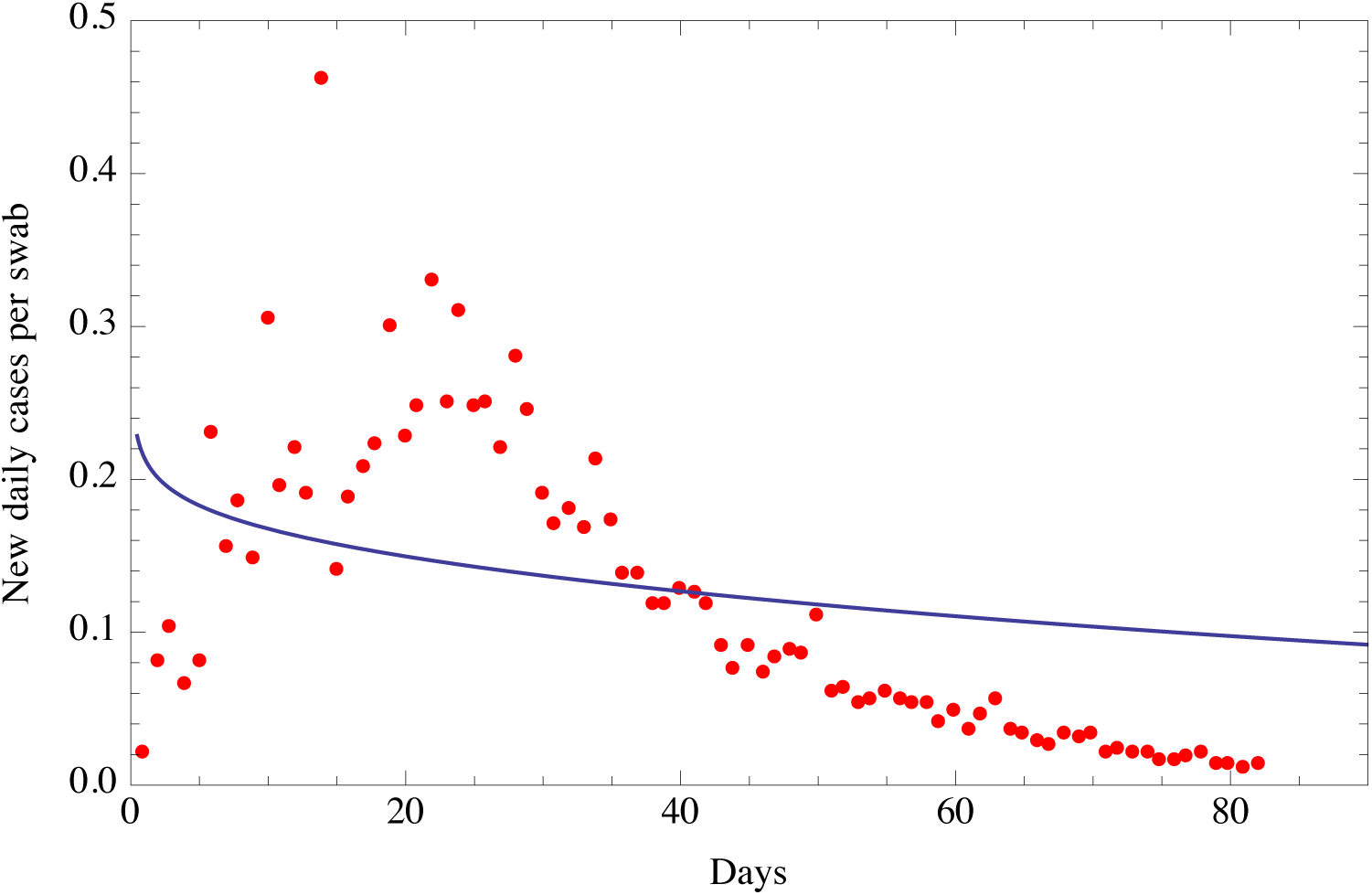
Fit of the ratio of the number of new daily Covid-19 detections in Italy via nasopharyngeal swab tests on a given day divided by the number of nasopharyngeal swab tests given in Italy that day. This fit is obtained with a function of the type of a Weibull distribution^4^, 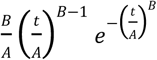, with two parameters *A* and *B*. The beginning date is February 25. Root Mean Square of the residuals is 0.0839

**Fig. 2e.**
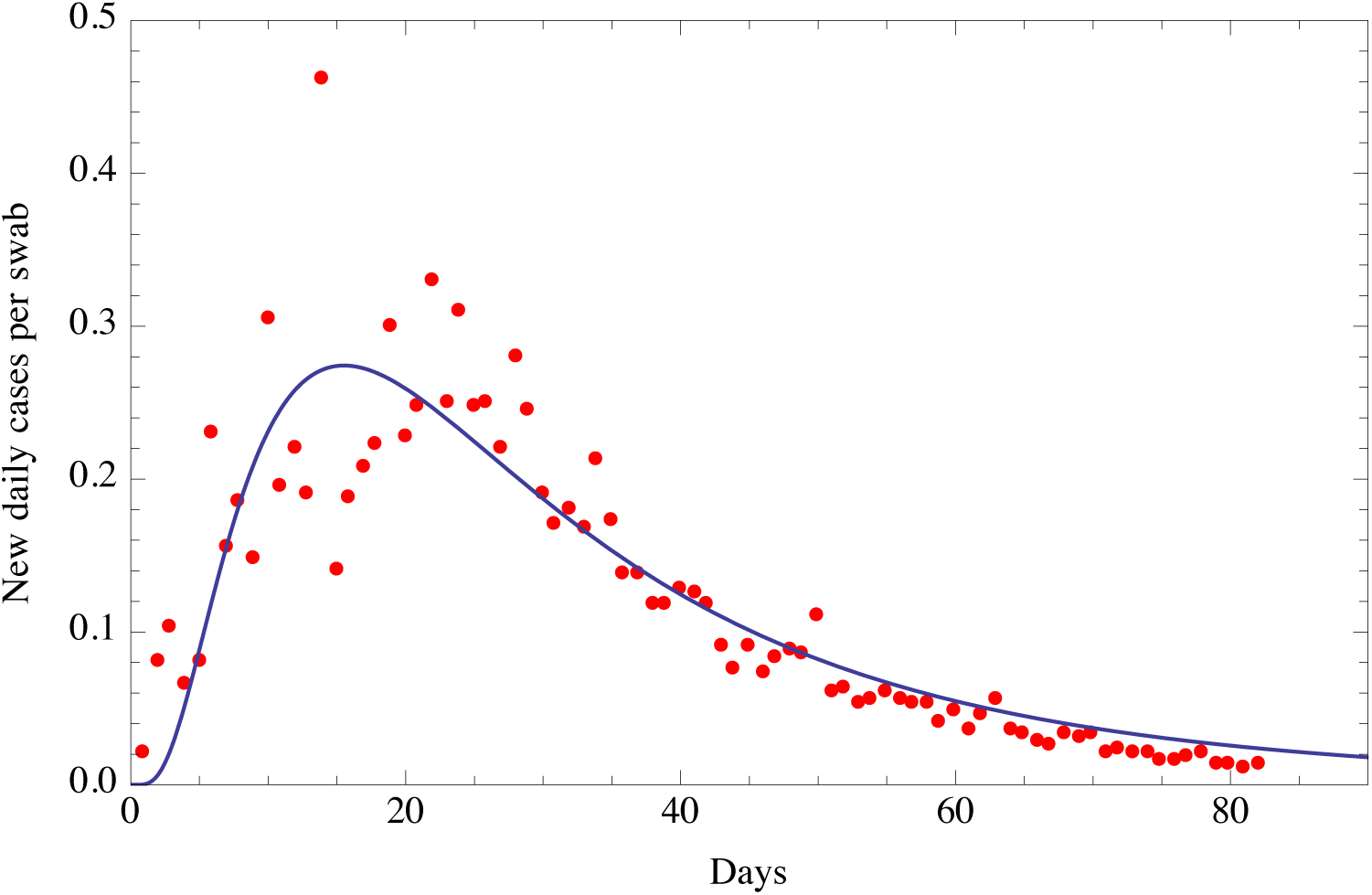
Fit of the ratio of the number of new daily Covid-19 detections in Italy via nasal swab tests on a given day divided by the number of nasal swab tests given in Italy that day. This fit is obtained with a function of the type of a Lognormal distribution^5^,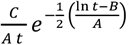, with three parameters *A, B* and *C*. The beginning date is February 25. Root Mean Square of the residuals is 0.0426

**Fig. 2f.**
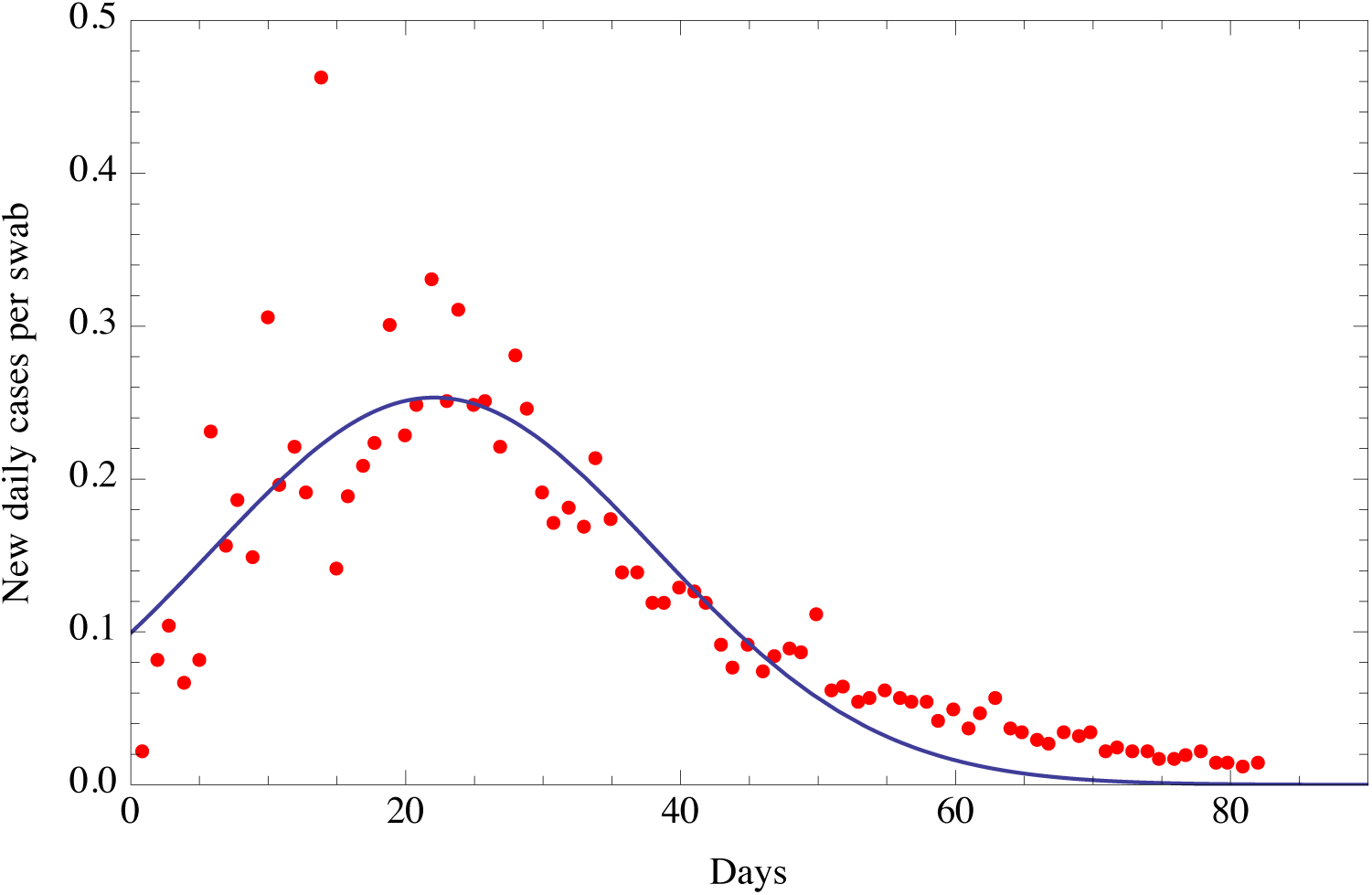
Fit of the ratio of the number of new daily Covid-19 detections in Italy via nasopharyngeal swab tests on a given day divided by the number of nasopharyngeal swab tests given in Italy that day. This fit is obtained with a function of the type of a Gauss function^3^, 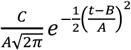, with three parameters *A, B* and *C*. The beginning date is February 25. Root Mean Square of the residuals is 0.0436

### 2.1 Prediction capabilities of the distributions fitting the data

As a relevant test to select the distribution which is more suitable to predict the evolution in time of the new daily cases per swab, we consider three different intervals of daily ratios: from February 25 until May 2, from February 25 until April 18 and from February 25 until April 4, i.e., two weeks before the date of May 16, four weeks before May 16 and six weeks before May 16, respectively. We then fit the daily ratios during these three periods with each of the seven distributions and finally test which one gives the best predictions for the measured ratios up to May 16, from May 2, from April 18 and from April 4, respectively. We test the predictions of each distribution by visual inspection (Table 1) and by calculating the lowest absolute value of the mean and Root Mean Square (RMS) of the residuals (difference between the measured daily data and the fitting function), as reported in Table 2. As shown in Tables 1 and 2, the distributions providing the best predictions are the Planck, Beta and Gamma distributions. The Weibull distribution does not fit well the data.

**Table 1.**
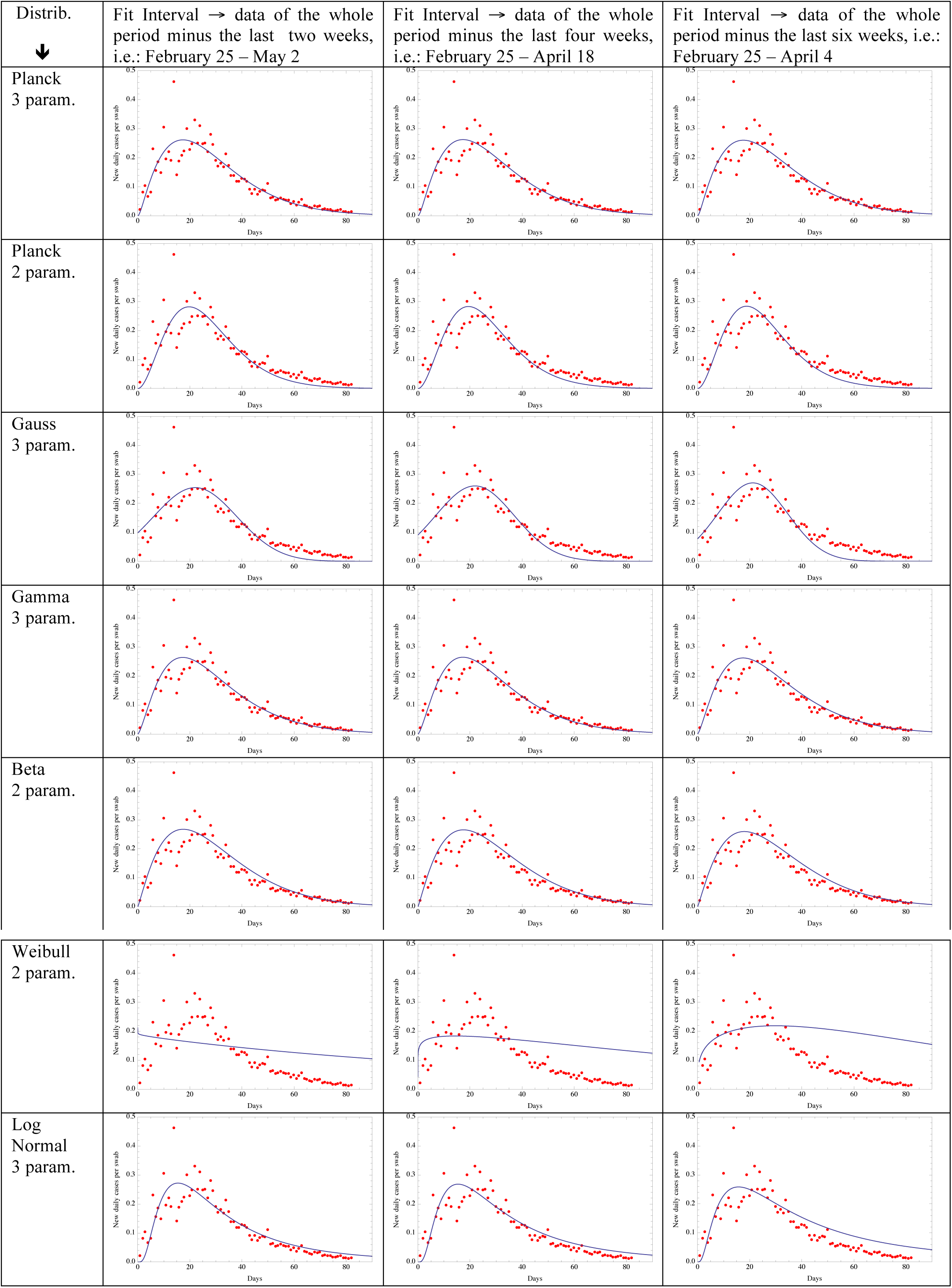
Plots of the seven distributions (rows) obtained by fitting the daily ratios over three different time intervals (columns): from February 25 up to two weeks before May 16, four weeks before May 16 and six weeks before May 16, respectively.

**Table 2.** Mean and Root Mean Square of the residuals calculated over the last two weeks, the last four weeks and the last six weeks (i.e., from May 2 April 18, April 4 up to May 16), corresponding to the seven fitting distributions.

| Distribution<br>↓ | Mean and RMS between<br>May 2 and May 16 | Mean and RMS between<br>April 18 and May 16 | Mean and RMS between<br>April 4 and May 16 |
| --- | --- | --- | --- |
| Planck 3 par. | -0.0044<br>0.0034 | -0.0045<br>0.0050 | 0.0034<br>0.0115 |
| Planck 2 par. | -0.0144<br>0.00499 | -0.0188<br>0.0069 | -0.0214<br>0.01097 |
| Gauss 3 par. | -0.0187<br>0.0061 | -0.0259<br>0.0095 | -0.0323<br>0.0148 |
| Gamma 3 par. | -0.0029<br>0.0034 | -0.0031<br>0.00499 | 0.0057<br>0.0111 |
| Beta 2 par | 0.00018<br>0.0031 | 0.0039<br>0.0062 | 0.01199<br>0.0133 |
| Weibull 2 par | 0.0951<br>0.0046 | 0.112<br>0.0086 | 0.1422<br>0.0199 |
| Lognormal 3 par | 0.0134<br>0.0034 | 0.0184<br>0.0048 | 0.0414<br>0.0098 |

## 3. Analysis of new daily cases per swab of Covid-19 in Italy and Monte Carlo simulations

In the present section, we first analyze the expected dates of a substantial reduction in the ratio of the daily diagnosed cases per swab in Italy and we then perform a Monte Carlo simulation with 25000 runs to evaluate the random uncertainties in these dates.

Using the best fitting distribution, i.e., a Planck distribution with three parameters (see previous section 2) and Gamma-type and Gauss-type distributions, we find the dates when we expect that the ratio of daily diagnosed cases per swab will be below certain thresholds. We choose these thresholds to be equal to the minimum measured ratio of cases per swab (in the range February 25 to May 4), equal to 0.02163, incidentally corresponding to the first available datum of such ratio occurred on February 25; one fifth of it and one tenth of it. These dates are summarized in Table 3.

**Table 3.** Dates of a substantial reduction in the ratio of cases per swab according to the three fitting distributions: Planck-type with three parameters, Gamma-type and Gauss-type.

|  | Threshold | Planck 3 param.<br>(days since Feb. 25)<br>date | Gamma 3 param.<br>(days since Feb. 25)<br>date | Gauss 3 param.<br>(days since Feb. 25)<br>date |
| --- | --- | --- | --- | --- |
| Historical minimum from February 25 to May 4 | 0.02163 | (71)<br>May 5 | (72)<br>May 6 | (58)<br>April 22 |
| 5 times smaller than the historical minimum | 0.004326 | (93)<br>May 27 | (96)<br>May 30 | (69)<br>May 3 |
| 10 times smaller than the historical minimum | 0.002163 | (102)<br>June 5 | (106)<br>June 9 | (72)<br>May 6 |

However, several uncertainties can influence the number of the new daily cases of the Covid-19 pandemic, in addition to the number of nasopharyngeal swabs that have increased with time as explained in the previous section. Another source of uncertainty is related to the fact that the number of swabs is associated both to new cases and to the repeated ones of persons already tested as positive. However if we assume that the percentage of re-tested people is approximately constant such error should be small.

To possibly estimate the uncertainties in the number of new daily cases per swab, we use Monte Carlo simulations^3,10,11^ with 25000 runs, similar to what was done in our previous works^1,2^, and the differences in the predictions among the best distributions considered previously. The Monte Carlo simulation should account mainly for random uncertainties. The uncertainty we consider in the Monte Carlo simulation does not take into account the difference between the real number of cases (which is unknown) and the daily diagnosed ones which can be one order of magnitude larger, or even more. However, it is usual in statistics to use a sample as being representative of the population under study and the present Monte Carlo simulation is performed for the number of diagnosed daily cases per swab.

For the convenience of the reader, we summarize here the procedure used previously^1,2^ for the total number of cases which is instead applied here to the ratio of cases per swab. The number of runs has been largely increased from 150 to 25000. We assume a measurement uncertainty in the daily number of diagnosed cases per swab equal to 20% of each daily ratio (Gaussian distributed). Then, a random matrix (*m×n*) is generated, where *n* (columns) is the number of observed days and *m* (rows) is the number of random outcomes, which we have chosen to be 25000. Each number in the matrix is part of a Gaussian distribution with mean equal to 1 and sigma equal to 0.2 (i.e., 20% of 1), either row-wise and column-wise. So, starting from the *n* nominal values of the daily data per swab, we generated *n* Gaussian distributions with 25000 outcomes, with mean equal to the *n* nominal values and with 20% standard deviation. Then, for each of the 25000 simulations, those *n* values (corresponding to the daily cases per swab of *n* days) are fitted with a three parameter function of the type of a Planck function (see section 2) and we then determine the date at which the number of new daily cases will be less than a certain threshold that, for example, we choose to be equal to the minimum measured ratio of cases per swab (in the range February 25 to May 4), equal to 0.02163, one fifth of it and one tenth of it.

Finally, we calculate the mean and the standard deviation of the 25000 simulations for these three cases. The results are reported in Table 4.

**Table 4.** Results of the Monte Carlo simulation with 25000 runs.

| Threshold description | Threshold | Planck type distribution with three parameters |  |
| --- | --- | --- | --- |
|  |  | Mean value of day number and corresponding date | Standard deviation (days) |
| Historical minimum from February 25 to May 4 | 0.02163 | (71)<br>May 5 | 1.6 |
| 5 times smaller than the historical minimum | 0.004326 | (93)<br>May 27 | 2.3 |
| 10 times smaller than the historical minimum | 0.002163 | (102)<br>June 5 | 2.9 |

In Fig. 3, is reported the histogram of the frequencies versus the day of a substantial reduction in the number of daily cases per swab corresponding to less than 0.004326, i.e., to less than one fifth of the minimum value of cases per swab measured during the period February 25 to May 4. The histogram approaches a Gaussian with mean equal to day 93, approximately corresponding to what reported in Fig. 1. The standard deviation is approximately 3 days.

**Fig. 3.**
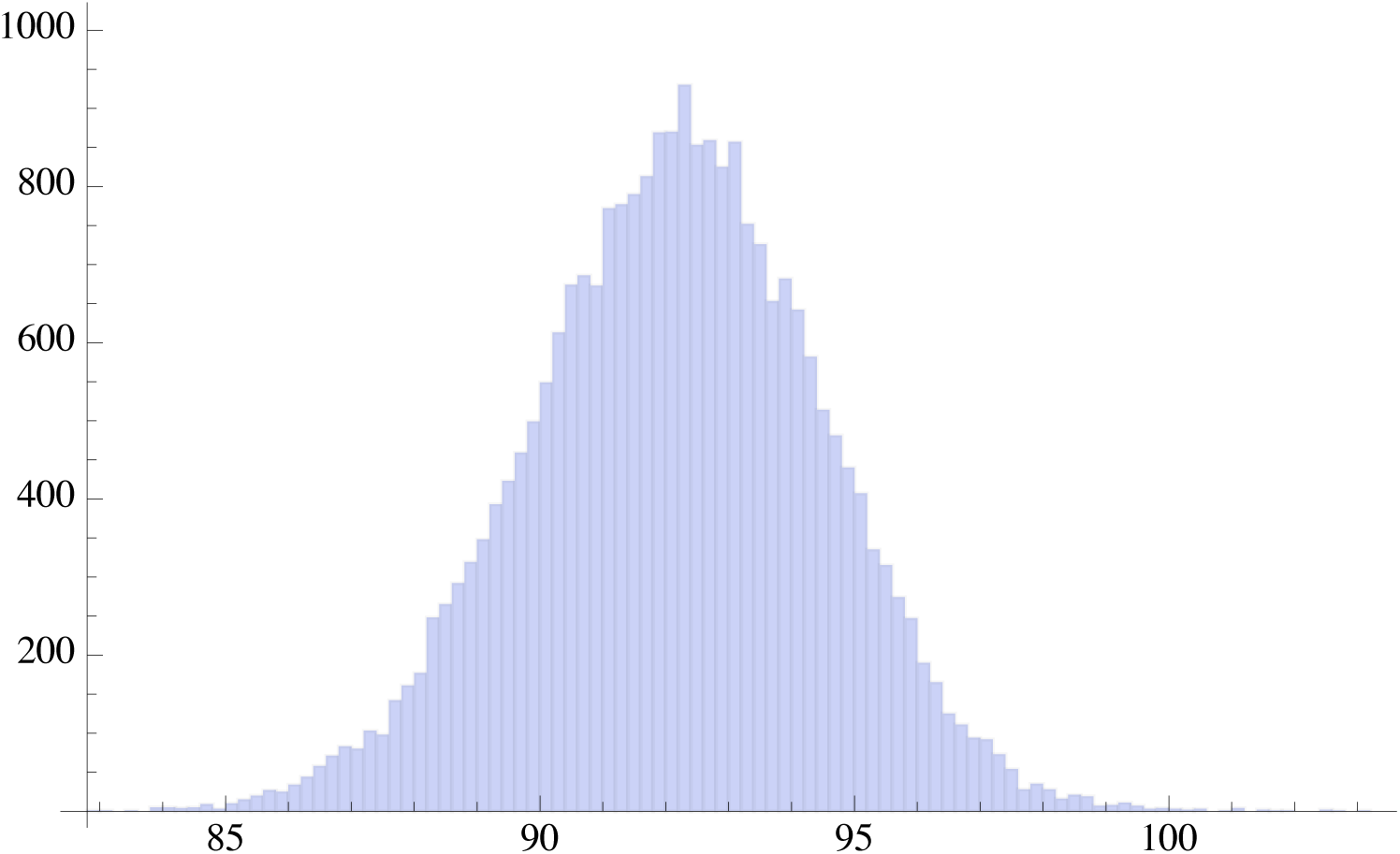
Histogram of the 25000 runs of the Monte Carlo simulation for Italy: frequencies versus the day in which a substantial reduction in the daily cases per swab is less than 0.004326, i.e., less than one fifth of the minimum value of cases per swab measured during the period February 25 to May 4.

### 3.1 Analysis of each region of Italy

We independently analyze each of the twenty Italian regions from February 25, 2020 (included) until April 25, 2020 (included). As in the previous sections also here we consider the ratio of daily cases over the number of daily swabs. Indeed, the number of daily swabs and other relevant conditions vary quite differently from one region to the other so that we can get an indication of the spread between different regions. To calculate such spread, we evaluate the date of the reduction of the daily cases per swab in each region below a certain threshold which is chosen to be the minimum value of cases per swab in each region in the range under study (25 February- 25 April). We then fit the daily ratios of each region using a function of the type of the Planck law with three parameters and finally, for each region, we obtain the date at which the number of daily cases per swab reduces below the given threshold for each region. In Fig. 4 we report the 20 dates corresponding to the 20 regions. The calculation of the spread of the 20 dates provides a 1-sigma standard deviation of 11.4 days.

**Fig. 4.**
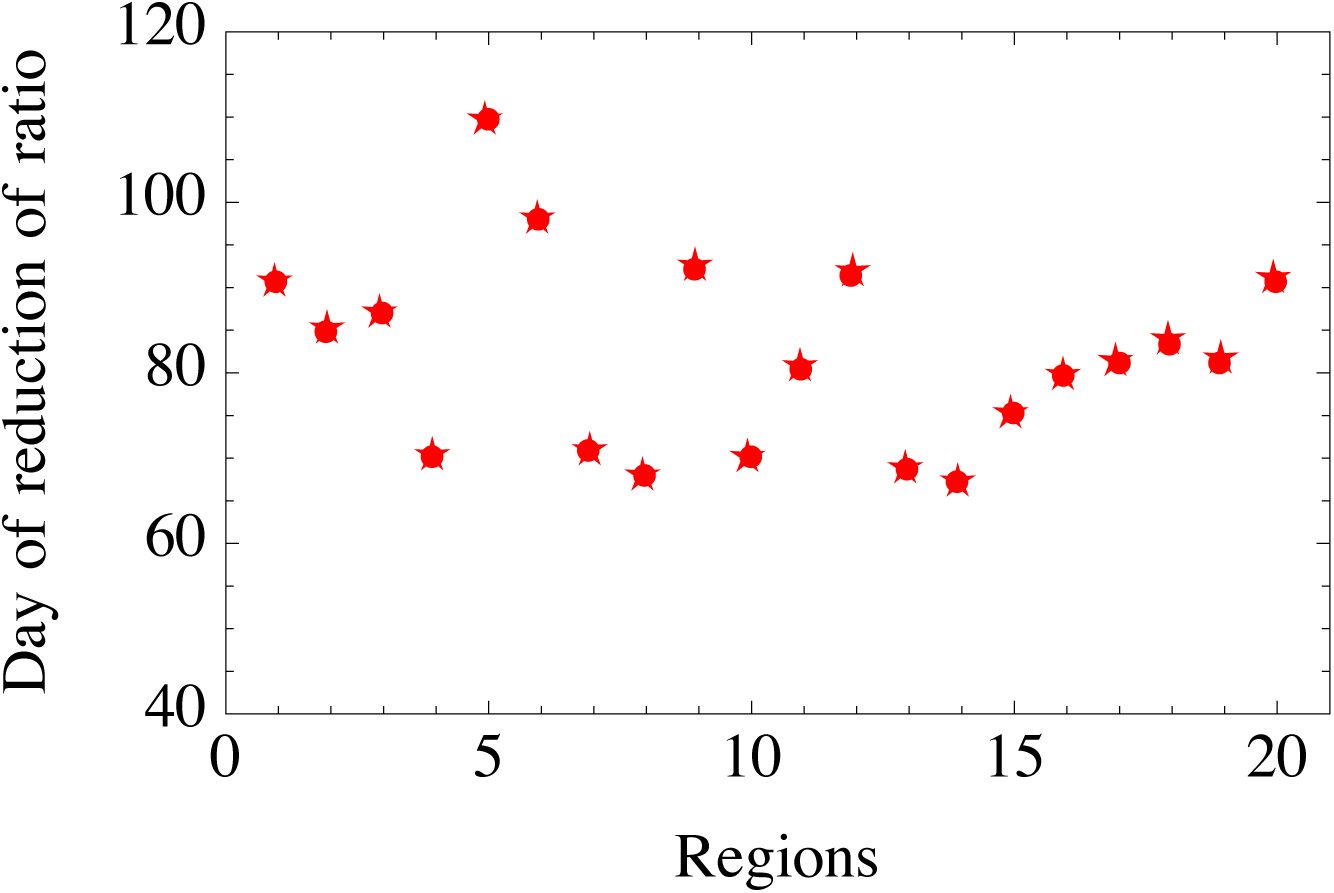
Days of a substantial reduction in the number of daily cases per swab for each of the 20 Italian regions. The 1-sigma standard deviation is 11.4 days

This result may also be displayed using the Central Limit Theorem. We choose a sample of 30 regions, with repetition, out of the 20 regions, for 1200000 times. We then calculate the mean of each sample and we report the frequencies of the mean of the 1200000 samples in the histogram of Fig. 5. We obtain a Gaussian with a standard deviation of 2.03 days, that, multiplied for the square root of 30, gives back a standard deviation of approximately 11.4 days as shown in Fig. 4. This represents the fact that each region has quite different conditions with regard to the intensity and phase of the pandemic, the number of daily swabs and other factors.

**Fig. 5.**
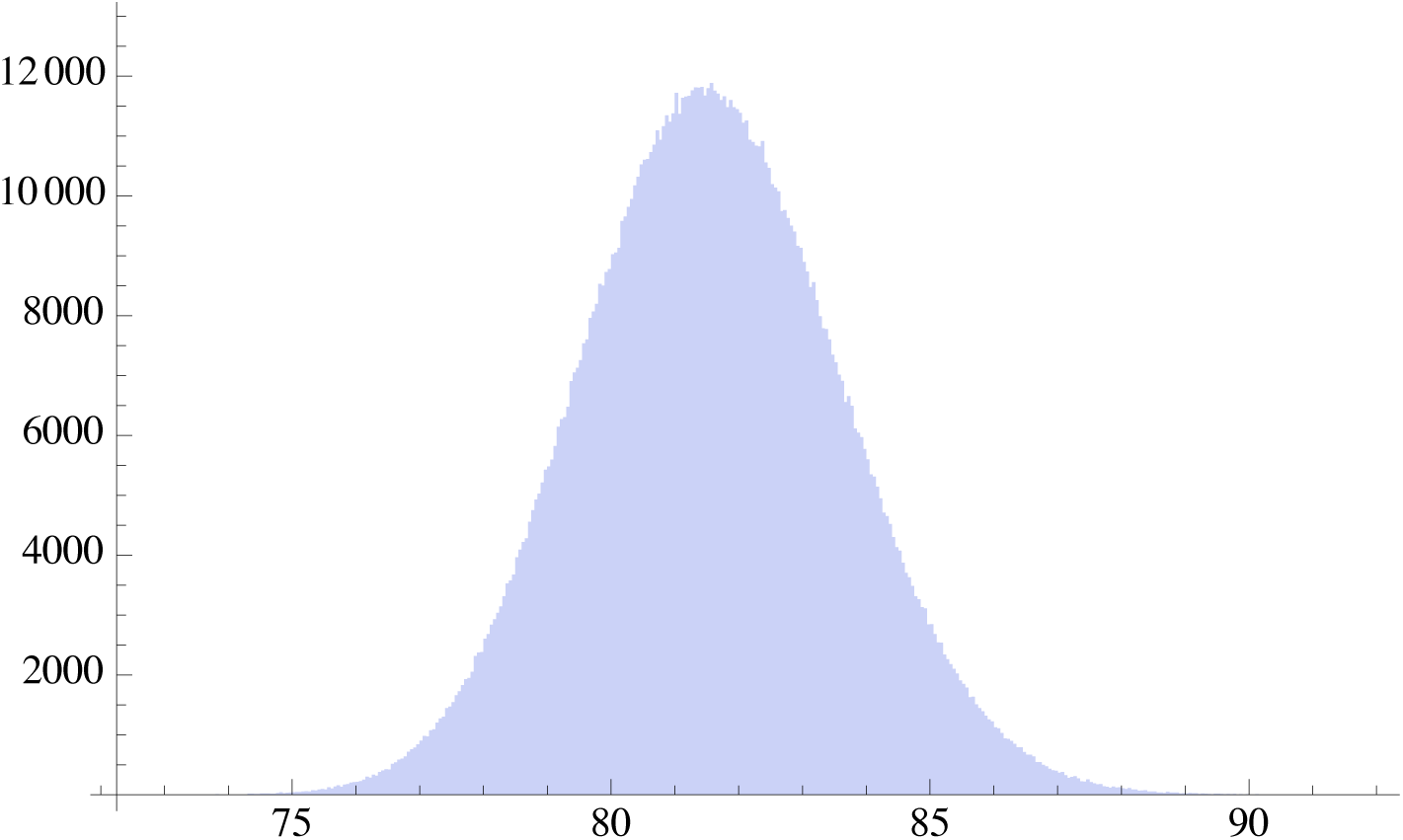
Result of 1200000 simulations using the 20 Italian regions. The figure is the histogram of the mean of each sample of the regions. The standard deviation is 11.13

## 4. Modelling the daily swabs

Since the actual number of new daily cases is much higher than the measured ones^12^, by increasing the number of daily swabs (which depends on factors that are difficult to predict, such as the daily availability of reagents and specialized personnel), the number of new daily cases would also increase. Therefore, since the number of daily swabs is significantly changing in time, the ratio of new daily cases per swab is more meaningful than the number of daily cases. Nevertheless, in this section we consider five possible trajectories for the daily number of swabs corresponding to some relevant situations. We also tried to model the number of daily swabs using a Gaussian distribution, a Planck distribution and a linear monotonic increasing distribution, however for the sake of brevity we do not report this part here. The five cases we consider are: 9000, i.e., about the number of daily swabs equal to the mean of daily swabs between February 15 and March 26 (the date of our first analysis^1,2^); 24000, i.e., about the mean between February 15 and April 25; 45000, i.e., about the mean between March 26 and April 25; 67000, i.e., about the maximum number of swabs up to April 25 and 100000, i.e., an estimated possible upper bound to the number of daily swabs. In Table 5 are reported those five cases for a Planck distribution and the Gamma distribution, i.e., the best distributions with regard to prediction capability as reported in Tables 1 and 2, compared to a Gauss distribution which gives an earliest bound for the predicted date of a substantial reduction in the number of daily cases.

**Table 5.** Day when the number of new daily cases becomes lower than a normalized threshold according to the predictions of the three distributions reported here. The beginning date is February 25 (included).

| Normalized threshold for the number of daily cases | Distribution→<br>Daily swabs ↓ | Day number and (date) with Gauss 3 parameters | Day number and (date) with Planck 3 parameters | Day number and (date) with Gamma 3 parameters |
| --- | --- | --- | --- | --- |
| 100 cases | **9000** equal to mean up to 26 March | 63<br>(27 April) | 80<br>(14 May) | 82<br>(16 May) |
| 267 cases | **24000** equal to mean up to 25 April |  |  |  |
| 500 cases | **45000** equal to mean from 27 Mar to 25 Apr |  |  |  |
| 744 cases | **67000** equal to max value up to 25 April |  |  |  |
| 1111 cases | **100000** upper bound |  |  |  |

In our previous work^1,2^, a substantial reduction of the daily cases was chosen to occur when the number of cases reduced to about 100. However, since the number of daily swabs is significantly changing and thus the number of cases increases as the number of swabs, we normalize 100 by multiplying it by the ratio: “number of swabs divided by 9000”. The number 9000, as mentioned earlier, is approximately the average number of swabs up to March 26. Table 5 shows the dates of a substantial reduction of new daily cases predicted with the three different distributions. The date of a substantial reduction of cases in the population should be independent of the number of swabs since it describes the real evolution of the pandemic at a certain stage. For this reason we introduce the normalization just described. In other words, the different numbers of daily cases reported in the first column of Table 5 would represent the same actual condition of the pandemic, i.e., 100 cases obtained with 9000 swabs would, e.g., be equal to 500 cases obtained with 45000 swabs.

Since the number of daily swabs was increasing after March 26 by a factor of more than 5.6 with respect to our previous analysis, a substantial decrease in the number of new daily cases is reached later with respect to our previous mathematical prediction^1,2^. As mentioned at the beginning of this section, a better indication of the evolution of the pandemic is provided by the behavior of new daily cases per swab as reported in Figs. 1 and 2 and in section 3. Indeed. the maximum value of the number of new daily cases per swab was 0.462 reached on March 9, whereas the minimum measured value (in the range February 25 to May 4) was equal to 0.02163. According to our previous work^1,2^, the pandemic should have significantly reduced during the end of April. Indeed, on May 1, the ratio of daily cases per swab was 0.0265 (i.e., nearly its measured value at the beginning of the pandemic in Italy) and on May 15 the ratio was even smaller, reaching the historical minimum (from February 25 to May 16) of 0.0116.

In Fig. 6 we report a 3D representation of the number of new daily cases as a function of the number of daily swabs, from 0 to 100000, and of the day, from February 25, using a Planck law distribution.

**Fig 6.**
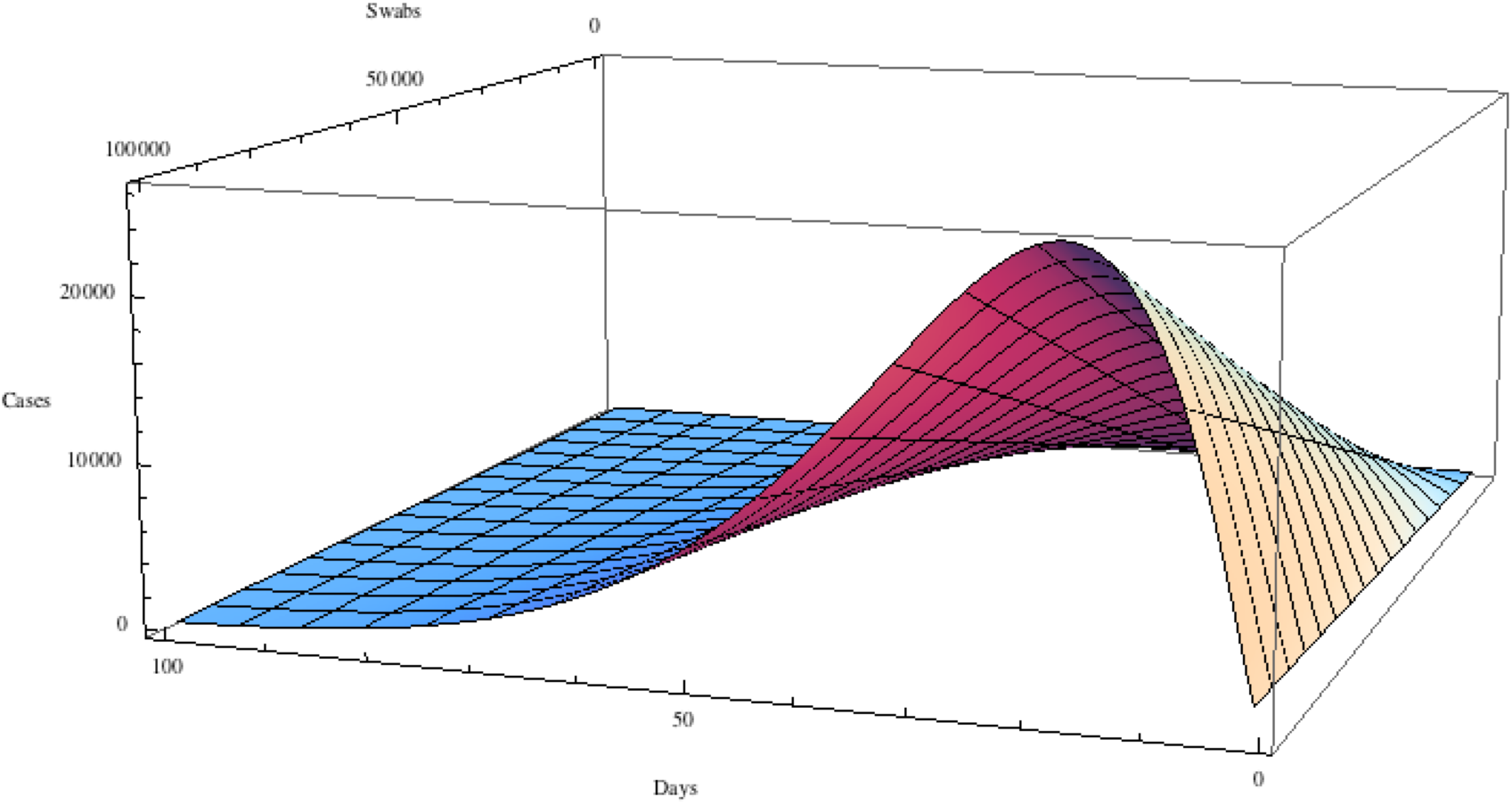
Three dimensional representation of the number of daily cases as a function of the number of daily swabs (from 0 to 100000) and of the day, from February 25, for a distribution of the type of Planck law with three parameters used to describe the new daily cases per swab.

## 5. Conclusions

Since the number of daily swabs was rapidly increasing in Italy after March 26, we fit the ratio of the new daily cases per swab using several functions, including the Gaussian, Weibull, Lognormal, Beta and Gamma distributions and a Planck law function.^1^ Indeed, using the best fitting distribution, i.e., the Planck distribution with three parameters, and depending on the chosen threshold for the daily cases per swab (historical minimum from February 25 to May 4, or one fifth of it, or one tenth of it), we found that a substantial reduction in the daily cases per swab will occur between May 5 and June 5.

Furthermore, considering that is practically impossible to predict the evolution of the number of daily swabs, we consider five possible relevant trajectories for the number of daily swabs. By considering these five trajectories and the Gauss, Gamma and Planck distributions, the range of dates for a substantial decrease in the number of new daily cases goes from April 27 (in agreement with our previous findings) to May 16, depending on the chosen distribution. To characterize a substantial decrease of the pandemic, we have assumed a threshold of 100 cases per day when an average of 9000 swabs per day are used. However, if for instance, the number of daily swabs is instead 67000 (the maximum number of swabs per day up to April 25), the corresponding indication of that substantial decrease in the pandemic is given by a much higher number of cases, i.e., 744. Indeed, since the actual number of new daily cases is much higher than the measured ones, by increasing the number of daily swabs the number of new daily cases will also increase.

To estimate the random uncertainty in the dates of a substantial reduction of the pandemic in Italy, we used a Monte Carlo simulation, with 25000 runs, which provides a 1-sigma random uncertainty of approximately three days (calculated for a threshold of one tenth of its historical minimum between February 25 and May 4). In addition, we also estimated the spread in a substantial reduction below a certain threshold of the daily cases per swab among the Italian regions.

## Data Availability

All data referred in the manuscript are publicily available. The sources are provided in the References of the paper

## Acknowledgments

We gratefully thank Richard Matzner (University of Texas at Austin) for helpful suggestions, Alessandro Paolozzi, and Claudio Paris (Centro Fermi).

## Footnotes

1 Incidentally this last one fits well the number of daily positive cases in China too.

